# Switching from febuxostat to dotinurad in patients with chronic kidney disease and hyperuricemia: a single-center, non-randomized study

**DOI:** 10.64898/2026.07.16.26358294

**Authors:** Taisuke Irifuku, Shigeharu Kashiwado, Takao Masaki

## Abstract

Recently, an observational study demonstrated that a lower fractional excretion of uric acid (FEUA) is significantly associated with a higher risk of kidney failure. This study aimed to assess the efficacy of switching from febuxostat to dotinurad, which increases FEUA, in patients with chronic kidney disease (CKD) and hyperuricemia (HUA). This was a non-randomized, open-label, single-center, prospective, single-arm study involving 60 patients with CKD and HUA who received febuxostat. Participants first underwent a 3-month observation period, followed by a 3-month intervention period, during which treatment was switched from febuxostat to dotinurad. The primary outcomes were changes from baseline to 3-months after switching in the estimated glomerular filtration rate (eGFR) calculated from serum creatinine (eGFRcreat) and serum cystatin C (eGFRcys), as well as the serum uric acid levels. The secondary outcome was defined as the correlation between ΔFEUA and the changes in both eGFRcreat(ΔeGFRcreat) and eGFRcys(ΔeGFRcys), respectively. During the observation period, mean eGFRcreat decreased significantly. The baseline eGFRcreat (mL/min/1.73 m²) was 36.0 ± 15.2, and the serum urate level (mg/dL) was 5.5 ± 1.2. During the intervention period, eGFRcreat increased in contrast to the significant decline observed in eGFRcys. After 3 months of switching to dotinurad, the mean serum UA levels increased significantly from 5.5 ± 1.2 to 6.1 ± 1.4 mg/dL, despite a significant elevation in FEUA. Both ΔeGFRcreat and ΔeGFRcys after switching to dotinurad were positively correlated with ΔFEUA. Switching from febuxostat to dotinurad resulted in discrepant changes in eGFRcreat and eGFRcys, suggesting that renal function should be assessed carefully after switch. Additionally, the risk of elevated serum UA levels should be considered when switching from febuxostat to dotinurad in patients with CKD.

## Introduction

Serum urate levels increase linearly with decreasing glomerular filtration rate owing to reduced excretion [1]. Therefore, hyperuricemia (HUA) is highly prevalent among patients with chronic kidney disease (CKD) [2]. Previous cohort studies have indicated that HUA is a potentially modifiable risk factor for the development and progression of CKD, cerebral and cardiovascular diseases, and mortality [3–7]. However, the superiority of urate-lowering therapy for improving renal outcomes remains controversial [8,9]. Single-center trials have shown that urate-lowering therapy with allopurinol or febuxostat can slow the progression of CKD over a short follow-up period [10,11]. In contrast, a febuxostat versus placebo randomized controlled trial on reduced renal function in patients with HUA complicated by CKD stage 3 demonstrated that febuxostat did not exert a suppressive effect on estimated glomerular filtration rate (eGFR) decline compared to placebo [12]. Therefore, urate-lowering treatment with febuxostat for HUA may not improve the renal prognosis.

Dotinurad is a novel uricosuric drug characterized as a selective urate reabsorption inhibitor that acts via selective inhibition of renal urate transporter 1 (URAT1), promoting uric acid (UA) excretion [13]. It has been evaluated for its non-inferiority to benzbromarone and febuxostat, and for its safety and efficacy in long-term use [14–16]. Recently, an observational study reported that lower fractional excretion of uric acid (FEUA) was associated with a higher risk of kidney failure [17], suggesting that accelerating UA excretion with dotinurad may be beneficial for kidney outcomes.

This study aimed to investigate the effectiveness of direct transition from febuxostat to dotinurad in patients with CKD and asymptomatic HUA. This study also aimed to determine whether this therapeutic approach can be implemented in routine clinical practice.

## Materials and Methods

### Study design, participants, and intervention

This single-arm, prospective, single-center, non-randomized, open-label study was conducted in a real-world clinical setting. The study participants were outpatients who visited the Division of Nephrology at the National Hospital Organization of Higashihiroshima Medical Center between April 1, 2024 and August 31, 2024. The main inclusion criteria for enrollment were patients with CKD stage G2–G4 (eGFR 15–89 mL/min/1.73 m ²) who received febuxostat for HUA at doses ranging from 10 to 40 mg/day. Patients with a history of active gouty arthritis were also excluded. This study was approved by the Ethics Committee of the National Hospital Organization of the Higashihiroshima Medical Center (approval No 2024-92), and informed consent was obtained in verbally from all patients who participated in the study. This study was registered in the University Hospital Medical Information Network (UMIN; 000060295).

The participants first underwent a 3-month observation period, followed by a 3-month dotinurad treatment period. Specifically, after a 3-month observation period before the intervention, febuxostat was discontinued, and dotinurad treatment (administered orally) was initiated and continued during the intervention period (3 months). In most cases, the dose of dotinurad was adjusted to achieve a serum UA level <6.0 mg/dL. In patients with serum UA levels >8.0 mg/dL despite receiving the maximum dose of dotinurad, febuxostat was administered. Other medications that could affect the eGFR and/or serum UA levels, including losartan (9 cases), loop diuretics (10 cases), thiazide diuretics (8 cases), and sodium glucose cotransporter 2 inhibitors (35 cases), did not change during the study period. Additionally, no participants were taking fenofibrate. The study design and conversion dose of dotinurad from febuxostat are shown in Figure 1.

**Fig. 1.**
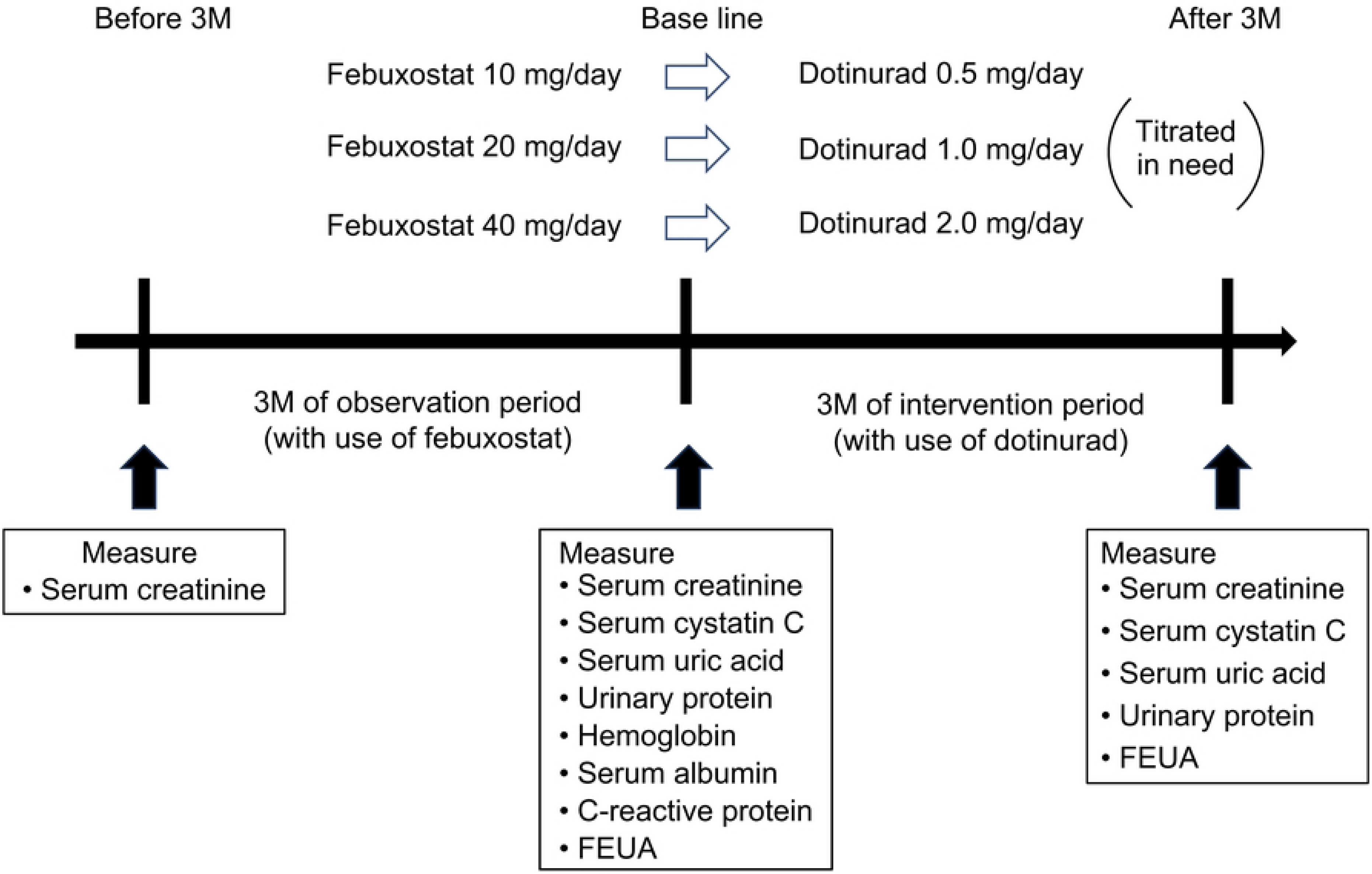
Schematic presentation of this study.

### Data collection

Demographic, clinical, and laboratory data were reviewed. Serum UA, serum creatinine (Cr), serum cystatin C (Cys), hemoglobin, serum albumin, hemoglobin A1c, C-reactive protein, urinary Cr, urinary protein, and urinary UA levels were measured at our hospital using standardized reagents and methods. The degree of proteinuria was quantified using the spot urine protein-to-creatinine ratio (g/g Cr). FEUA was also calculated from spot urine samples before dotinurad administration and after 3 months.

Renal function was expressed as eGFR calculated from serum Cr (eGFRcreat) and serum Cys (eGFRcys), which were determined using the formula proposed by the Japanese Society of Nephrology [18,19].

Data on adverse events that occurred after the start of dotinurad administration were collected.

### Study Outcome

The primary outcomes were changes from baseline to 3-months after switching in the eGFRcreat, eGFRcys, and serum UA levels. The secondary outcome was defined as the correlation between ΔFEUA and both ΔeGFRcreat and ΔeGFRcys, respectively.

### Statistical analysis

Categorical data are presented as numbers and percentages. Continuous data were reported as the mean ± standard deviation when normally distributed and as the median and interquartile range when not normally distributed. Normality was assessed by visual inspection of histograms and by calculating skewness and kurtosis. For paired comparisons, the distribution of within-subject changes was primarily considered. Within-group comparisons were performed using the paired *t*-test for normally distributed data and the Wilcoxon signed-rank test for non-normally distributed data. P-values were corrected using the Bonferroni method to avoid multiple tests.

The changes in eGFR from 3 months before administration to baseline (ΔeGFR-pre) and from baseline to 3 months after administration (ΔeGFR-post) were compared using the paired *t*-test. Stratified analyses were performed according to sex, age, CKD stage, and primary disease status.

All statistical analyses were performed using SPSS version 29.0.2.0 Windows; IBM Japan, Tokyo, Japan. A p-value of ≤0.05 was considered statistically significant.

## Results

Informed consent was obtained from 63 patients. After excluding one patient who withdrew consent and two who did not meet the eligibility criteria, 60 patients received the intervention, and all of them completed the 3-month intervention period. The baseline characteristics and laboratory data of the participants are presented in Table 1. At the start of the intervention, the mean age was 65.2 ± 15.2 years, and 70% of the patients were male. The mean eGFRcreat was 36.0 ± 15.2 mL/min/1.73 m², and the mean serum UA level was 5.5 ± 1.2 mg/dL. Of the 60 participants, four were classified as having CKD stage G2, 28 as having stage G3, and 28 as having stage G4. None of the participants reported any adverse effects from dotinurad during the intervention period.

**Table 1.** Patients’ profile at baseline.

|  | **data (n=60)** | **male (n=42)** | **female (n=18)** |
| --- | --- | --- | --- |
| **age** | 65.2 ± 15.2 | 62.7 ± 16 | 70.9 ± 10.6 |
| **sex** |  |  |  |
| **male** | 42, 70.00% |  |  |
| **female** | 18, 30.00% |  |  |
| **body weight (kg)** | 66.7 ± 15.4 | 70.2 ± 15.8 | 58.544 ± 10.1 |
| **systolic blood pressure (mmHg)** | 127.4 ± 11.4 | 127.8 ± 11.7 | 126.5 ± 10.3 |
| **diastolic blood pressure (mmHg)** | 71.8 ± 11.1 | 72.5 ± 11.5 | 70.1 ± 9.1 |
| **primary disease** |  |  |  |
| **chronic glomerulonephritis** | 17, 28.30% | 10, 23.80% | 7, 38.90% |
| **diabetic nephropathy** | 14, 23.30% | 9, 21.40% | 5, 27.80% |
| **nephrosclerosis** | 18, 30.00% | 13, 30.90% | 5, 27.80% |
| **other** | 11, 18.30% | 10, 23.80% | 1 , 0.055 |
| **hemoglobin (g/dL)** | 12.9 ± 1.7 | 13.4 ± 1.6 | 11.8 ± 1.1 |
| **serum albumin (g/dL)** | 4.1 ± 0.5 | 4.1 ± 0.5 | 4.1 ± 0.3 |
| **HbA1c (%)** | 6 ± 0.7 | 6 ± 0.7 | 6.1 ± 0.7 |
| **C-reactive protein (mg/dL)** | 0.06 [0.02, 0.17] | 0.05, [0.02, 0.17] | 0.05, [0.02, 0.12] |
| **serum creatinine (mg/dL)** | $1.7 \pm 0.6$ | $1.7 \pm 0.6$ | $1.5 \pm 0.6$ |
| **eGFR creat (ml/min/1.73m<sup>2</sup>)** | $36 \pm 15.2$ | $38.3 \pm 15.6$ | $30.7 \pm 12.1$ |
| **cystatin C (mg/L)** | $2 \pm 0.7$ | $1.9 \pm 0.7$ | $2.3 \pm 0.7$ |
| **eGFR cys (ml/min/1.73m<sup>2</sup>)** | $36.6 \pm 20.3$ | $41 \pm 21.5$ | $26.2 \pm 10.7$ |
| **serum uric acid (mg/dL)** | $5.5 \pm 1.2$ | $5.7 \pm 1.1$ | $4.8 \pm 1$ |
| **urinary protein (g/gCr)** | 0.27, [0.10, 1.35] | 0.23, [0.09, 1.03] | 0.43, [0.19, 1.62] |
| **fractional excretion of uric acid (%)** | $6.7 \pm 2.6$ | $6.2 \pm 2.4$ | $7.9 \pm 2.7$ |

data presentation : n, % ; mean ± sd ; median [IQR].

Figure 2 presents a comparison of eGFRcreat values 3 months before, at baseline, and 3 months after the intervention. During the 3-month observation period, the mean eGFRcreat declined significantly from 37.8 ± 15.7 to 36.0 ± 15.2 mL/min/1.73 m², suggesting that renal dysfunction was progressive in these patients. In contrast, during the 3-month intervention period, the mean eGFRcreat increased from 36.0 ± 15.2 to 38.1 ± 16.9 mL/min/1.73 m². Accordingly, the mean ΔeGFRcreat-pre showed a negative value (-1.7 ± 3.1 mL/min/1.73 m²), whereas the mean ΔeGFRcreat-post showed a positive value (2.1 ± 3.7 mL/min/1.73 m²), and the difference between them was statistically significant. These effects of conversion to dotinurad on eGFRcreat were consistently observed regardless of sex, age, CKD stage at the start of the intervention or primary cause of CKD; however, no significant difference was found in the subgroup with diabetic nephropathy as the primary disease (Table 2).

**Fig. 2.**
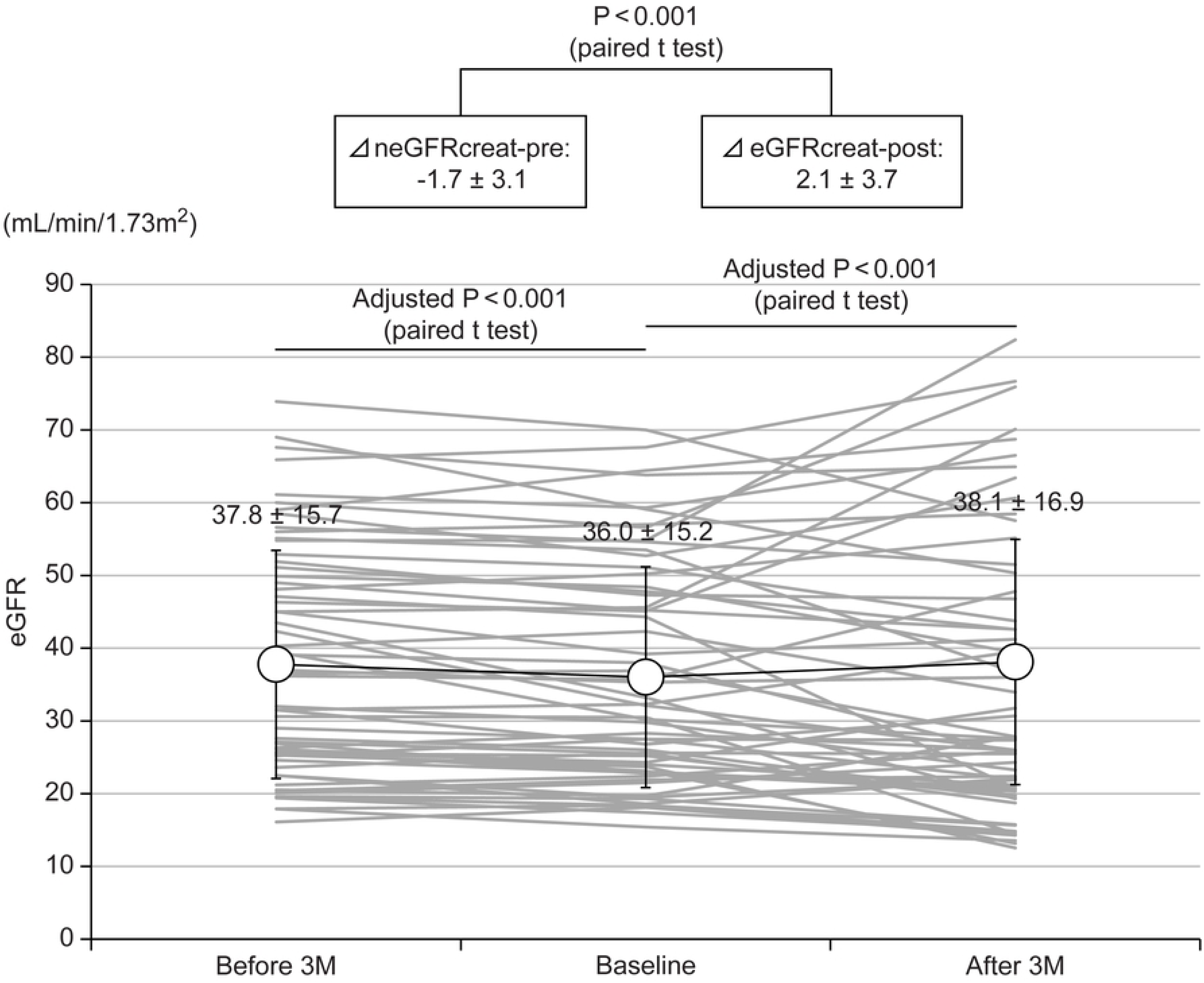
Change in eGFRcreat during the observation and intervention periods.

**Table 2.** Stratified analysis: Change in eGFRcreat before and after switching to dotinurad.

|  | n | ΔeGFR-pre: | ΔeGFR-post: | P-value |
| --- | --- | --- | --- | --- |
| **All** | 60 | -1.7 ± 3.1 | 2.1 ± 3.7 | **<0.001** |
| **gender** |  |  |  |  |
| **male** | 42 | -1.7 ± 3.5 | 2.1 ± 4 | **0.001** |
| **female** | 18 | -2 ± 1.9 | 2.1 ± 2.8 | **<0.001** |
| **age** |  |  |  |  |
| **65 or higher** | 36 | -2.1 ± 3.1 | 2.5 ± 3.7 | **<0.001** |
| **< 65** | 24 | -1.3 ± 3.1 | 1.4 ± 3.6 | **0.024** |
| **CKD stage at base line** |  |  |  |  |
| **G2** | 4 | -0.2 ± 4.5 | 1.9 ± 4.4 | 0.66 |
| **G3** | 28 | $-2.7 \pm 3.6$ | $3.5 \pm 4.2$ | **<0.001** |
| **G4** | 28 | $-1 \pm 2$ | $0.7 \pm 2.3$ | **0.013** |
| **primary disease** |  |  |  |  |
| **chronic glomerulonephritis** | 17 | $-1.6 \pm 2.9$ | $2.5 \pm 4.4$ | **0.022** |
| **diabetic nephropathy** | 14 | $-1.2 \pm 3.2$ | $1.6 \pm 2.6$ | 0.069 |
| **nephrosclerosis** | 18 | $-2.9 \pm 3.7$ | $2.3 \pm 4.1$ | **0.006** |
| **other** | 11 | $-0.8 \pm 1.9$ | $1.8 \pm 3.1$ | **0.015** |

data presentation: mean ± sd.
P-value: paired t test.

The changes in various parameters, including eGFRcys, before and after switching to dotinurad (at baseline and after 3 months) are shown in Table 3. After 3 months of switching to dotinurad, the mean serum UA levels increased significantly from 5.5 ± 1.2 to 6.1 ± 1.4 mg/dL in patients with CKD stage G2–G4, despite a significant elevation in FEUA. Ten of the 60 participants required the addition of febuxostat during the intervention period because their serum UA levels increased to ≥8 mg/dL. These patients tended to have lower baseline eGFR (eGFRcreat 24.4 ± 6.1mL/min/1.73 m²) and a higher prevalence of proteinuria (2.5 ± 3.5 g/gCr), suggesting that the efficacy of dotinurad might be reduced in patients with more advanced CKD. The primary outcome analyzed by excluding patients who required concomitant febuxostat is presented in Supplementary Table S1. Interestingly, although the mean eGFRcreat increased significantly in the intervention period, as previously demonstrated, switching from febuxostat to dotinurad resulted in a significant decrease in eGFRcys from 36.6 ± 20.3 to 35.0 ± 19.0 mL/min/1.73 m² (Figure 3). These findings indicate a potential interference of dotinurad and/or febuxostat with renal tubular Cr secretion, rather than a true improvement in renal function following their administration.

**Fig. 3.**
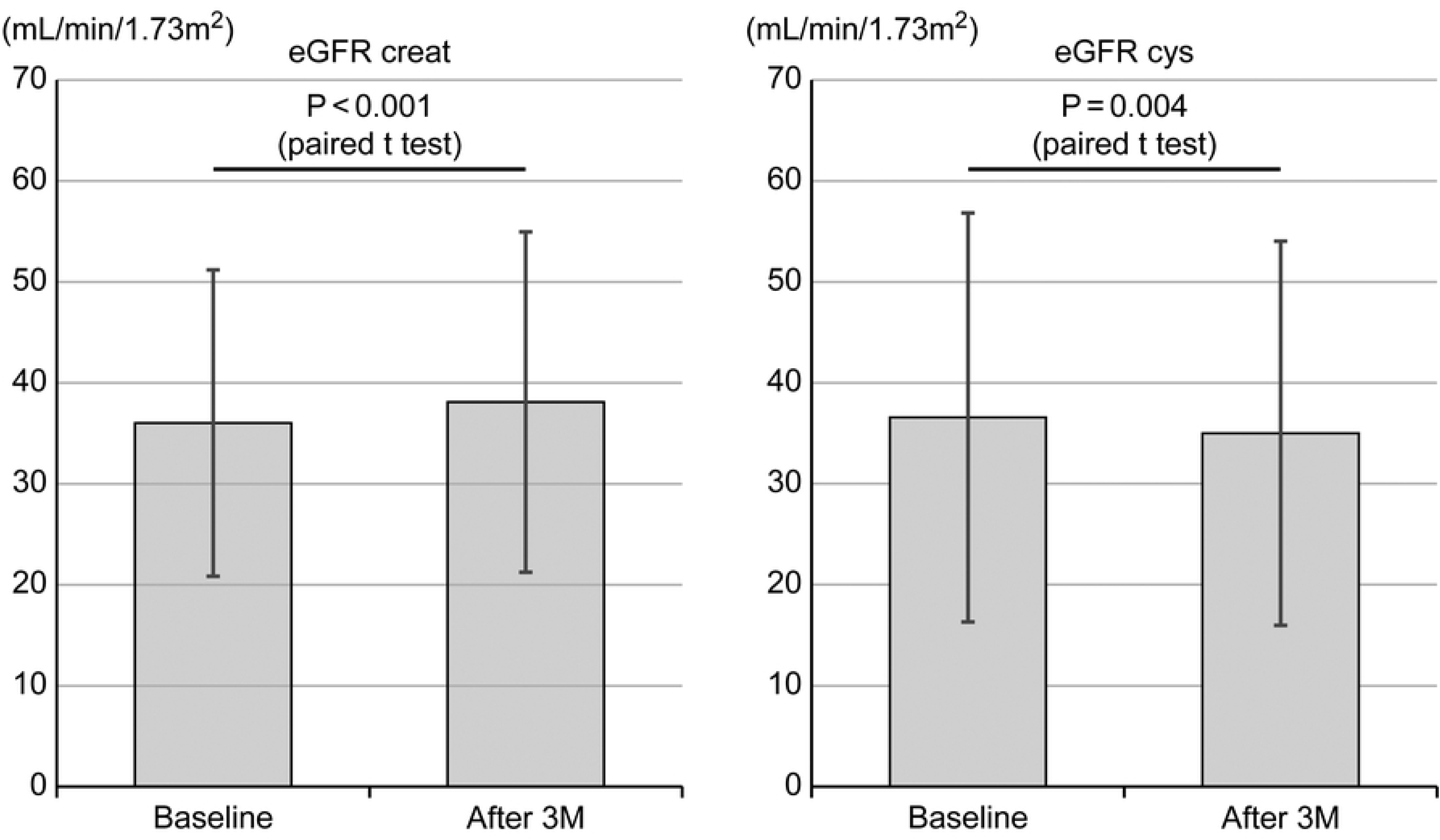
Changes in eGFRcreat and eGFRcys during 3 months of intervention.

**Table 3.** Change in various parameters before and after switching dotinurad.

|  | **Baseline** | **After 3M** | **P-value** |  |
| --- | --- | --- | --- | --- |
| **serum creatinine (mg/dL)** | 1.7 ± 0.6 | 1.6 ± 0.6 | **0.007** | a |
| **eGFR creat (ml/min/1.73m<sup>2</sup>)** | 36 ± 15.2 | 38.1 ± 16.9 | **<0.001** | a |
| **cystatin C (mg/L)** | 2 ± 0.7 | 2.1 ± 0.7 | **0.019** | a |
| **eGFR cys (ml/min/1.73m<sup>2</sup>)** | 36.6 ± 20.3 | 35 ± 19 | **0.004** | a |
| **serum uric acid (mg/dL)** | 5.5 ± 1.2 | 6.1 ± 1.4 | **0.001** | a |
| **urinary protein (g/gCr)** | 0.27, [0.10, 1.35] | 0.27, [0.08, 1.26] | 0.18 | b |
| **fractional excretion of uric acid (%)** | $6.7 \pm 2.6$ | $10.8 \pm 4.8$ | **<0.001** | **a** |

data presentation: mean ± sd: median [IQR].
P-value: a, paired t test; b, Wilcoxon signed-rank test.

Figure 4 presents the relationship between the increase in ΔFEUA and both ΔeGFRcreat-post and ΔeGFRcys-post during the 3-month intervention period, respectively. Significant positive correlations were observed, particularly with eGFRcys.

**Fig. 4.**
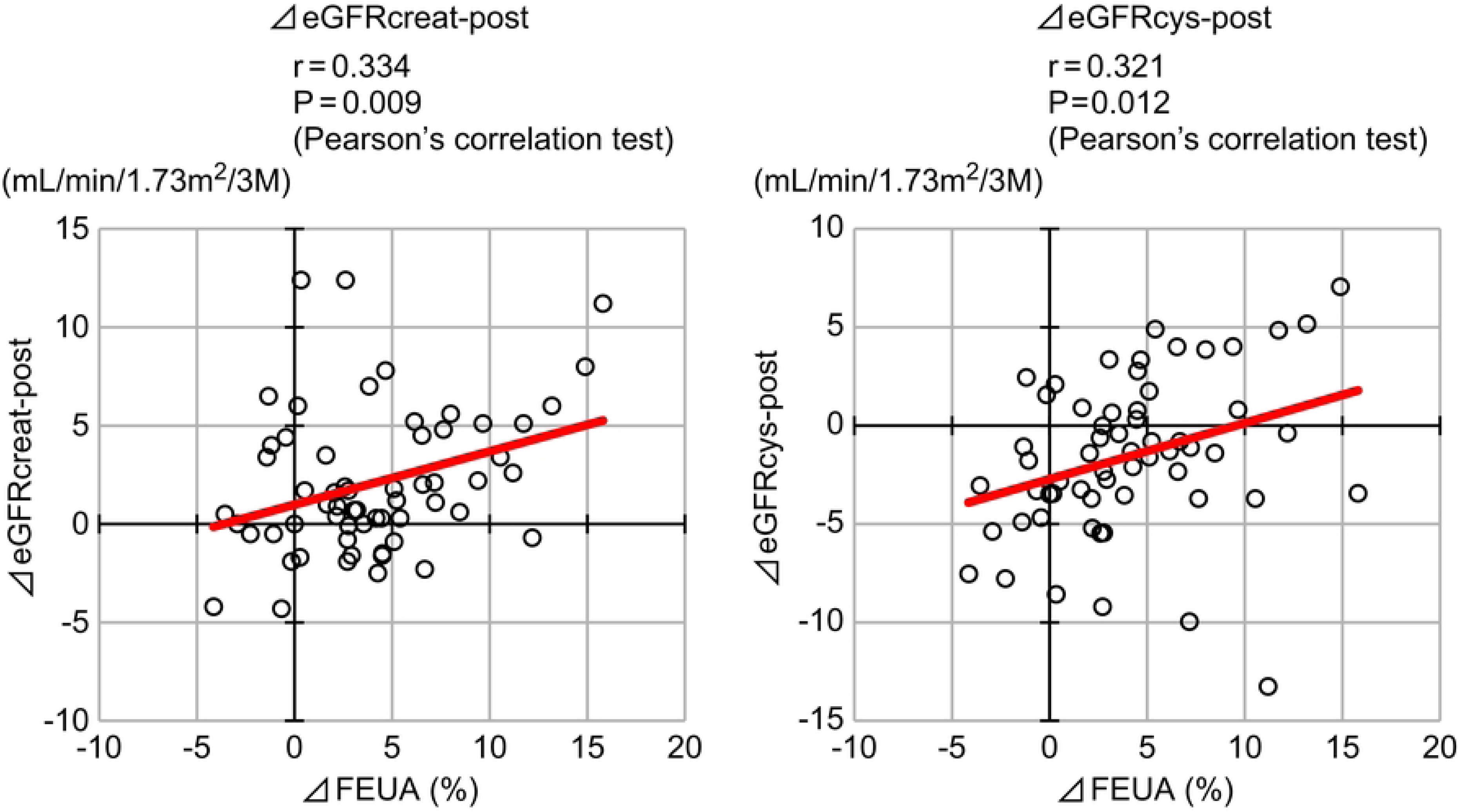
Relationship between the increase in ΔFEUA and both ΔeGFRcreat-post and ΔeGFRcys-post after switching to dotinurad.

## Discussion

In this single-center study of patients with CKD and HUA, the effectiveness of switching from febuxostat to dotinurad over 3 months was prospectively investigated. The major finding was that this change in treatment not only led to an improvement in eGFRcreat, but also resulted in a significant decrease in eGFRcys. This switch also resulted in a mean increase in serum UA of 0.6 mg/dL. Additionally, a clear relationship was observed between increased FEUA and both ΔeGFRcreat-post and ΔeGFRcys-post values. These findings suggest that both febuxostat and the selective URAT1 inhibitor dotinurad might affect Cr secretion in renal tubules and that renal function should be assessed carefully after switch. Additionally, the risk of elevated serum UA levels should be considered when switching from febuxostat to dotinurad in patients with CKD.

Dotinurad has been assessed for its non-inferiority to febuxostat in terms of long-term efficacy in patients without CKD [15,16]. However, in a single-dose pharmacokinetic study of dotinurad in patients with CKD stages G2–G3, serum UA reduction was attenuated in patients with CKD stage G3 compared to that in patients with CKD stage G1 [20]. This study performed drug transitions based on switching doses established for patients without CKD. Nevertheless, the mean serum UA level significantly increased from 5.5 ± 1.2 to 6.1 ± 1.4 mg/dL. Therefore, the risk of elevated serum UA levels should be considered when switching from febuxostat to dotinurad in patients with severe renal impairment. Although our study showed a decline in eGFRcys after switching to dotinurad, a retrospective observational study reported that dotinurad administration improved eGFRcys [21]. In this observational study, multivariate analysis revealed that the decrease in serum UA levels after dotinurad administration was only significant factor influencing the improvement of renal function. In the present study, an increase in serum UA level was observed after switching from febuxostat to dotinurad in CKD patients. The elevation of serum UA may explain the eGFRcys changes contrary to the previous observational study; however, further studies are needed to clarify the short-term relationship between eGFRcys and serum UA levels.

Notably, as shown in Figure 3, switching from febuxostat to dotinurad in patients with CKD resulted in a paradoxical response between eGFRcreat and eGFRcys. Furthermore, ΔeGFRcreat-pre during the observation period was generally comparable to ΔeGFRcys-post during the intervention period. This suggests that renal function may have consistently declined throughout the study period. Serum Cr concentration is a widely used biomarker of kidney function; however, it can be elevated for reasons unrelated to a reduced GFR (pseudoacute kidney injury [AKI]) [22]. Several antibacterial, antiviral, antifungal, antiparasitic agents, fibrates, and corticosteroids are associated with pseudo-AKI. The underlying mechanisms are multiple and diverse, ranging from interference with the serum Cr measurement method to inhibition of renal tubular secretion of Cr via various transporters [23]. Cys levels can help to distinguish between true and pseudo-AKI cases. On the other hand, it is known that Cys levels are affected by thyroid function and systemic inflammation; therefore, care must be taken as it is not Discrepancies between eGFRcreat and eGFRcys are frequently encountered in clinical practice. Because such discrepancies carry a risk of drug overdose, particularly for medications with a narrow therapeutic window, it is essential to carefully determine which index is more accurate by considering the patient’s muscle mass and comorbidities. a flawless biomarker [24,25].

Previous studies have reported that organic anion transporter 3 (OAT3) expressed on the basolateral membrane of renal tubular cells, other transporters such as OAT2, organic cation transporter 2 (OCT2), OCT3 on the apical membrane and, multidrug and toxin extrusion protein 1 (MATE1) and MATE2K on the basolateral membrane are also involved in renal tubular Cr secretion [26,27]. To the best of our knowledge, it has only been reported that febuxostat and its major acyl-glucuronide metabolite inhibit OAT3 in human embryonic kidney 293 cells via competitive and non-competitive mechanisms [28]. Additionally, dotinurad has been reported not to inhibit OAT3 [29]. Therefore, we hypothesized that switching from febuxostat to dotinurad promoted OAT3-mediated tubular secretion of Cr, resulting in a short-term increase in eGFRcreat.

Despite concerns that increased urinary UA excretion aggravates kidney damage by promoting UA crystallization, a cohort study revealed that lower FEUA levels were associated with a higher risk of kidney failure even in a subgroup of patients with aciduria [17]. This is consistent with the positive correlation observed between increased FEUA and eGFRcys-post levels in our study.

This study has several limitations. First, it is unclear whether the findings could be generalized to other ethnic or age groups because the subjects analyzed were all outpatients of a single institute. Second, this study is single-arm observational design and cannot distinguish treatment effects from natural disease progression, seasonal variations, or regression to the mean. The observed eGFRcreat decline during the observation period and subsequent increase during intervention cannot be definitively attributed to the drug switch. Third, because this study targeted outpatients and FEUA was calculated using spot urine samples, it may be less accurate than assessments based on 24-hour urine collection. Fourth, due to health insurance restrictions, repeated measurements of Cys were difficult, resulting in missing eGFRcys data during the observation period. It is difficult to determine whether the post-intervention decline of Cys represents accelerated progression or continuation of baseline trends. This study’s findings should be interpreted with caution because of the difficulty in controlling for selection bias and patient backgrounds. To overcome these limitations, long-term randomized controlled trials comparing dotinurad with other standard therapies in a larger number of patients are required.

## Conclusions

In conclusion, these findings imply that, when transitioning from febuxostat to dotinurad in patients with CKD and HUA, the observed improvement in eGFRcreat must be interpreted carefully, potentially because of the effect of the drug on tubular Cr secretion. Additionally, the risk of elevated serum UA levels should be considered after switching from febuxostat to dotinurad in patients with CKD.

## Data Availability

The data presented in this study are available on request from the corresponding author.

## Acknowledgment

None.

## Supporting information

**Supplementary Table S1. Change in various parameters before and after switching dotinurad among patients not receiving febuxostat**

## References

1. Jing J, Kielstein JT, Schultheiss UT, Sitter T, Titze SI, Schaeffner ES, et al. Prevalence and correlates of gout in a large cohort of patients with chronic kidney disease: the German Chronic Kidney Disease (GCKD) study. Nephrol Dial Transplant. 2015;30: 613–621. doi: 10.1093/ndt/gfu352.

2. Juraschek SP, Kovell LC, Miller ER, Gelber AC. Association of kidney disease with prevalent gout in the United States in 1988-1994 and 2007-2010. Semin Arthritis Rheum. 2013;42: 551–561. doi: 10.1016/j.semarthrit.2012.09.009.

3. McDonagh TA, Metra M, Adamo M, Gardner RS, Baumbach A, Böhm M, et al. 2021 ESC Guidelines for the diagnosis and treatment of acute and chronic heart failure: developed by the Task Force for the diagnosis and treatment of acute and chronic heart failure of the European Society of Cardiology (ESC) with the special contribution of the Heart Failure Association (HFA) of the ESC. Rev Esp Cardiol (Engl Ed). 2022;75: 523. doi: 10.1016/j.rec.2022.05.005.

4. Li M, Hu X, Fan Y, Li K, Zhang X, Hou W, et al. Hyperuricemia and the risk for coronary heart disease morbidity and mortality a systematic review and dose-response meta-analysis. Sci Rep. 2016;6: 19520. doi: 10.1038/srep19520.

5. Kim SY, Guevara JP, Kim KM, Choi HK, Heitjan DF, Albert DA. Hyperuricemia and coronary heart disease: a systematic review and meta-analysis. Arthritis Care Res (Hoboken). 2010;62: 170–180. doi: 10.1002/acr.20065.

6. Kojima S, Sakamoto T, Ishihara M, Kimura K, Miyazaki S, Yamagishi M, et al. Prognostic usefulness of serum uric acid after acute myocardial infarction (the Japanese Acute Coronary Syndrome Study). Am J Cardiol. 2005;96: 489–495. doi: 10.1016/j.amjcard.2005.04.007.

7. Bose B, Badve SV, Hiremath SS, Boudville N, Brown FG, Cass A, et al. Effects of uric acid-lowering therapy on renal outcomes: a systematic review and meta-analysis. Nephrol Dial Transplant. 2014;29: 406–413. doi: 10.1093/ndt/gft378.

8. Badve SV, Pascoe EM, Tiku A, Boudville N, Brown FG, Cass A, et al. Effects of allopurinol on the progression of chronic kidney disease. N Engl J Med. 2020;382: 2504–2513. doi: 10.1056/NEJMoa1915833.

9. Goicoechea M, de Vinuesa SG, Verdalles U, Ruiz-Caro C, Ampuero J, Rincón A, et al. Effect of allopurinol in chronic kidney disease progression and cardiovascular risk. Clin J Am Soc Nephrol. 2010;5: 1388–1393. doi: 10.2215/CJN.01580210.

10. Siu YP, Leung KT, Tong MKH, Kwan TH. Use of allopurinol in slowing the progression of renal disease through its ability to lower serum uric acid level. Am J Kidney Dis. 2006;47: 51–59. doi: 10.1053/j.ajkd.2005.10.006.

11. Sircar D, Chatterjee S, Waikhom R, Golay V, Raychaudhury A, Chatterjee S, et al. Efficacy of febuxostat for slowing the GFR decline in patients with CKD and asymptomatic hyperuricemia: a 6-month, double-blind, randomized, placebo-controlled trial. Am J Kidney Dis. 2015;66: 945–950. doi: 10.1053/j.ajkd.2015.05.017.

12. Kimura K, Hosoya T, Uchida S, Inaba M, Makino H, Maruyama S, et al. Febuxostat therapy for patients with stage 3 CKD and asymptomatic hyperuricemia: a randomized trial. Am J Kidney Dis. 2018;72: 798–810. doi: 10.1053/j.ajkd.2018.06.028.

13. Ishikawa T, Takahashi T, Taniguchi T, Hosoya T. Dotinurad: a novel selective urate reabsorption inhibitor for the treatment of hyperuricemia and gout. Expert Opin Pharmacother. 2021;22: 1397–1406. doi: 10.1080/14656566.2021.1918102.

14. Hosoya T, Sano T, Sasaki T, Fushimi M, Ohashi T. Dotinurad versus benzbromarone in Japanese hyperuricemic patient with or without gout: a randomized, double-blind, parallel-group, phase 3 study. Clin Exp Nephrol. 2020;24 Suppl 1: 62–70. doi: 10.1007/s10157-020-01849-0.

15. Hosoya T, Furuno K, Kanda S. A non-inferiority study of the novel selective urate reabsorption inhibitor dotinurad versus febuxostat in hyperuricemic patients with or without gout. Clin Exp Nephrol. 2020;24 Suppl 1: 71–79. doi: 10.1007/s10157-020-01851-6.

16. Hosoya T, Fushimi M, Okui D, Sasaki T, Ohashi T. Open-label study of long-term administration of dotinurad in Japanese hyperuricemic patients with or without gout. Clin Exp Nephrol. 2020;24 Suppl 1: 80–91. doi: 10.1007/s10157-019-01831-5.

17. Asahina Y, Sakaguchi Y, Oka T, Hattori K, Kawaoka T, Doi Y, et al. Association between urinary uric acid excretion and kidney outcome in patients with CKD. Sci Rep. 2024;14: 5119. doi: 10.1038/s41598-024-55809-9.

18. Matsuo S, Imai E, Horio M, Yasuda Y, Tomita K, Nitta K, et al. Revised equations for estimated GFR from serum creatinine in Japan. Am J Kidney Dis. 2009;53: 982–992. doi: 10.1053/j.ajkd.2008.12.034.

19. Horio M, Imai E, Yasuda Y, Watanabe T, Matsuo S, Collaborators Developing the Japanese Equation for Estimated GFR. GFR estimation using standardized serum cystatin C in Japan. Am J Kidney Dis. 2013;61: 197–203. doi: 10.1053/j.ajkd.2012.07.007.

20. Fukase H, Okui D, Sasaki T, Fushimi M, Ohashi T, Hosoya T. Effects of mild and moderate renal dysfunction on pharmacokinetics, pharmacodynamics, and safety of dotinurad: a novel selective urate reabsorption inhibitor. Clin Exp Nephrol. 2020;24 Suppl 1: 17–24. doi: 10.1007/s10157-019-01825-3.

21. Takata T, Taniguchi S, Mae Y, Kageyama K, Fujino Y, Iyama T, et al. Comparative assessment of the effects of dotinurad and febuxostat on the renal function in chronic kidney disease patients with hyperuricemia. Sci Rep. 2025;15: 8990. doi: 10.1038/s41598-025-94020-2.

22. Zhang WR, Parikh CR. Biomarkers of acute and chronic kidney disease. Annu Rev Physiol. 2019;81: 309–333. doi: 10.1146/annurev-physiol-020518-114605.

23. Van Regemorter E, Ponlot E, Boland L, Gillion V, Devresse A. Drug-induced rise in serum creatinine: cystatin C to the rescue? Evidence, pitfalls and knowledge gaps. Eur J Intern Med. 2025;144: 106592. doi: 10.1016/j.ejim.2025.106592.

24. Wiesli P, Schwegler B, Spinas GA, Schmid C. Serum cystatin C is sensitive to small changes in thyroid function. Clin Chim Acta. 2003;338: 87–90. doi: 10.1016/j.cccn.2003.07.022.

25. Stevens LA, Schmid CH, Greene T, Li L, Beck GJ, Joffe MM, et al. Factors other than glomerular filtration rate affect serum cystatin C levels. Kidney Int. 2009;75: 652–660. doi: 10.1038/ki.2008.638.

26. Vallon V, Eraly SA, Rao SR, Gerasimova M, Rose M, Nagle M, et al. A role for the organic anion transporter OAT3 in renal creatinine secretion in mice. Am J Physiol Ren Physiol. 2012;302: F1293–F1299. doi: 10.1152/ajprenal.00013.2012.

27. Chu X, Bleasby K, Chan GH, Nunes I, Evers R. The complexities of interpreting reversible elevated serum creatinine levels in drug development: does a correlation with inhibition of renal transporters exist? Drug Metab Dispos. 2016;44: 1498–1509. doi: 10.1124/dmd.115.067694.

28. Tang LWT, Cheong TWH, Chan ECY. Febuxostat and its major acyl glucuronide metabolite are potent inhibitors of organic anion transporter 3: implications for drug-drug interactions with Rivaroxaban. Biopharm Drug Dispos. 2022;43: 57–65. doi: 10.1002/bdd.2310.

29. Yanai H, Adachi H, Hakoshima M, Iida S, Katsuyama H. A possible therapeutic application of the selective inhibitor of urate Transporter 1, dotinurad, for metabolic syndrome, chronic kidney disease, and cardiovascular disease. Cells. 2024;13: 450. doi: 10.3390/cells13050450.

